# Handheld chromatic pupillometry can reliably detect functional glaucomatous damage in eyes with high myopia

**DOI:** 10.1101/2022.11.27.22282795

**Authors:** Maxwell T. Finkelstein, Monisha E. Nongpiur, Rahat Husain, Shamira A. Perera, Mani Baskaran, Tina T Wong, Tin Aung, Dan Milea, Raymond P. Najjar

## Abstract

**Purpose:** To assess pupillary light responses (PLRs) in eyes with high myopia and evaluate the ability of handheld chromatic pupillometry (HCP) to identify glaucomatous functional loss in eyes with high myopia.

**Design:** Cross-sectional study with prospective data collection.

**Methods:** Participants included 28 emmetropes (EM), 24 high myopes without glaucoma (HM), and 17 high myopes with confirmed glaucoma (HMG), recruited at the Singapore National Eye Center. Monocular PLRs were evaluated using a custom-built handheld pupillometer that recorded changes in horizontal pupil radius in response to 9 seconds of exponentially increasing blue (469.1nm) and red (640.1nm) lights. Fifteen pupillometric features were compared between groups. A logistic regression model (LRM) was used to distinguish HMG eyes from non-glaucomatous eyes (EM and HM).

**Results:** All pupillometric features were similar between EM and HM groups. Phasic constriction to blue (P<0.001) and red (P=0.006) lights, and maximum constriction to blue light (P<0.001) were reduced in HMG compared to EM and HM. Pupillometric features of melanopsin function (PIPR AUC 0-12s (P<0.001) and PIPR 6s (P=0.01) to blue light) were reduced in HMG. Using only three pupillometric features, the LRM could classify glaucomatous from non-glaucomatous eyes with an AUC of 0.89 (95%CI: 0.77–1.00), sensitivity 94.1% (82.4%–100.0%) and specificity 78.8% (67.3%–90.4%).

**Conclusions:** PLRs to ramping-up light stimuli are unaltered in highly myopic eyes without other diagnosed ocular conditions. Conversely, HCP can distinguish glaucomatous functional loss in highly myopic eyes and may be a useful tool to detect/confirm the presence of glaucoma in patients with high myopia.

## Introduction

Myopia is the leading cause of vision impairment worldwide.^1^ This condition has reached epidemic magnitudes in East Asian countries, with a prevalence ranging from 20% to over 80%.^2–5^ Western populations are increasingly affected,^6,7^ and the global prevalence of myopia is expected to reach 50% by 2050 with almost one billion individuals having high myopia.^8^ High myopia, defined as a spherical equivalent less than or equal to -5.00 D, is associated with increased axial length,^9^ but also with retinal nerve fiber layer (RNFL) thinning,^10–14^ decreased visual field mean deviation (VFMD),^10,15–19^ and fundal abnormalities such as tilted discs and peripapillary atrophy.^20^ Furthermore, high myopia is a risk factor for other potentially blinding conditions, such as myopic macular degeneration, cataract, and glaucoma.^9^

Glaucoma diagnosis relies on ophthalmic clinical examination and tonometry as well as structural and functional investigations that include optical coherence tomography (OCT) imaging of the optic disc and RNFL, and standard automated perimetry (SAP).^21^ However, morphological changes in highly myopic eyes can render OCT imaging unreliable,^10,15–17^ and may elicit visual field abnormalities that mimic defects that are classically found in glaucoma.^19^ Both high myopia and glaucoma are highly prevalent in Asian populations,^2,4,22–24^ where over 70% of glaucoma patients are undiagnosed, even in developed countries like Singapore.^25^ Given the limitations of OCT and SAP, alternate methods for detecting glaucoma in high myopes must be investigated.

Amongst other functional ocular assessment tools like electroretinography (ERG), chromatic pupillometry has emerged as a non-invasive and objective method for evaluating photoreceptor integrity through the pupillary response to chromatic light stimuli. The pupillary light response (PLR) is triggered by intrinsically photosensitive retinal ganglion cells (ipRGCs) which express the photopigment melanopsin (λ_max_ ∼ 480nm) and integrate extrinsic inputs from rods and cones.^26–29^ As these cells elicit specific responses to different wavelengths of light, chromatic light stimulations have shown promise in differentially assessing the integrity of the inner and outer retina, as well as detecting glaucoma, even at an early stage.^30–37^

While studies using ERG have investigated large populations of high myopes and reported decreased amplitudes of a- and b-wave responses,^9^ studies using chromatic pupillometry have largely not found any effects of myopia on the PLR.^38–42^ Recently, however, Mutti and colleagues reported that myopic subjects displayed reduced constriction and faster redilation compared to emmetropes in response to 0.1 Hz blue light pulses.^43^ Nevertheless, all of the aforementioned studies investigated fewer than 10 high myopes, and used desktop devices for assessing the PLR. Moreover, the effect of glaucoma on the PLRs of highly myopic eyes remains undetermined.

In this cross-sectional study, we aimed to evaluate the impact of high myopia without visual field defects on the PLR, and tested the capability of handheld chromatic pupillometry (HCP) to distinguish functional glaucomatous damage in eyes with high myopia.

## Methods

### Participants

Seventy-three participants took part in this cross-sectional study, consisting of 24 highly myopic eyes (HM) without other ocular conditions, 21 highly myopic eyes with glaucoma (HMG), and 28 healthy emmetropic eyes (EM). Patients with unilateral or bilateral glaucoma were recruited from the glaucoma clinics of the Singapore National Eye Centre, while healthy participants without glaucoma, with or without high myopia, were recruited from the general outpatient clinics. Glaucomatous high myopes were diagnosed by fellowship trained glaucoma specialists based on the presence of glaucomatous optic neuropathy (loss of the neuroretinal rim and/or notching with nerve fiber layer defects attributable to glaucoma) accompanied by compatible visual field defects, with diagnosis confirmed via longitudinal evaluation of glaucomatous structural and functional defects over four years of follow-up. Non-glaucomatous participants were excluded if they had any family history of glaucoma, or any previous or existing ophthalmic health conditions (aside from refractive error in non-glaucomatous high myopes). High myopia was defined as spherical equivalent (SE) of -5.00 D or worse (less), while emmetropia was defined as SE between -1.50 D and +1.50 D.

All participants underwent a comprehensive ophthalmic examination including slit-lamp and gonioscopy examinations, best corrected visual acuity (LogMAR chart, Lighthouse International, New York, NY, USA), color vision testing (Ishihara plates, Kanehara & Co., Tokyo, Japan), automated refraction (Canon RK 5 Auto Ref-Keratometer, Canon, Tochigiken, Japan), IOP assessment via Goldmann applanation tonometry, and retinal nerve fiber layer imaging using high definition OCT (Cirrus version 6.0, Carl Zeiss Meditec, Dublin, CA, USA). If OCT signal strength was below 6, imaging was repeated up to twice more and the most reliable test result was used, even if it did not reach this threshold (n = 1 in the HMG group; n = 2 in the HM group). The visual fields of participants were assessed using standard automated perimetry (Humphrey visual field analyzer II model 750, Carl Zeiss Meditec, Dublin, CA, USA) with near refractive correction using the 24-2 Swedish Interactive Thresholding Algorithm (SITA Fast) and stimulus size III. Visual field assessment was repeated if false positive or false negative rates exceeded 33%, or if fixation loss rates exceeded than 20%. Questionnaires were used to collect demographic information, as well as medication and smoking history.

Participants were excluded if they were found to have additional ophthalmic conditions such as other optic neuropathies, retinopathies, ocular motor disorders, or pupillary abnormalities besides relative afferent pupillary defects in the glaucoma group. In addition, participants with intraocular lenses or dense cataracts (worse than NS2+) were excluded, as were those on medications that could affect the PLR and patients who had underwent intraocular surgery within the past 6 months. Participants with a known clinical diagnosis of psychiatric or neurological disorders (e.g. dementia), or diabetes mellitus were also excluded. The study was approved by the SingHealth Centralized Institutional Review Board, and written informed consent was obtained from all participants. Research procedures adhered to ethical principles outlined in the Declaration of Helsinki.

### Handheld Chromatic Pupillometry

Pupillometry testing took place in dimly lit rooms (< 1 lux) at the Singapore National Eye Center or Singapore Eye Research Institute clinics. The PLRs were assessed in all participants using a custom-built handheld chromatic pupillometer as described in Najjar et al.^37^ The study eye of each participant was recorded using an infra-red camera which was pre-focused on the pupil, and positioned approximately 60 degrees below the lower eyelid to prevent minor ptosis from affecting pupil size measurements. A tablet (iPad Mini, Apple, Cupertino, CA, USA) with a custom application was used to control the pupillometer, with this application allowing for real-time assessment of fixation and pupil detection. Participants were instructed to focus on a dim red central fixation zone (<0.1 lux) within the pupillometer. If a participant failed to maintain fixation or blinked excessively, the test was repeated.

Pupillometry testing was conducted for each participant using a 1 minute/eye procedure consisting of 10 seconds of darkness to measure baseline pupil size, followed by 9 seconds of exposure to exponentially increasing blue light (11.7 to 14.4 Log photons/cm^2^/s; λ_max_ = 469.1nm, full width at half maximum (FWHM) = 33nm), then 22 seconds of darkness to assess pupillary redilation before exposure to 9 seconds of exponentially increasing red light (11.9 to 14.3 Log photons/cm^2^/s ; λ_max_ = 640.1nm, FWHM = 17nm), and finally 10 seconds of darkness to assess redilation (Figure 1). Light exposure was monocular and administered to the study eye, with participants covering their fellow eye using their hand. Pupil size was measured as the pupil’s horizontal radius, and only one eye from each participant was included in the study. For participants with two eligible eyes, the study eye was chosen randomly.

**Figure 1.**
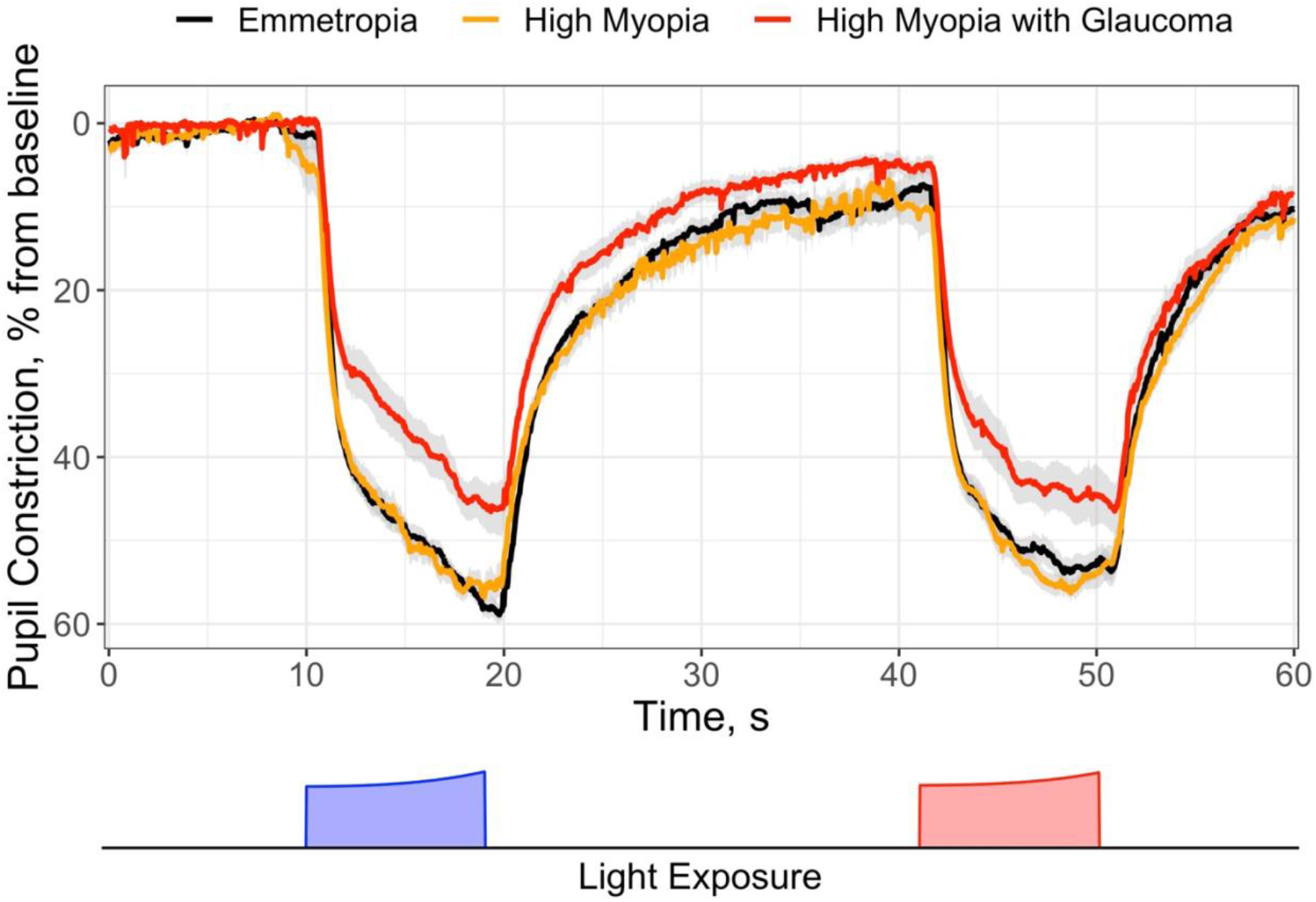
Mean baseline-adjusted pupillary light responses (PLRs) in emmetropes (n=28, black line), high myopes (n=24, orange line), and high myopes with glaucoma (n=17, red line). Pupillometry testing comprised 10 seconds of darkness to measure baseline pupil size, followed by 9 seconds of exposure to exponentially increasing blue light (11.7 to 14.4 Log photons/cm^2^/s; λ_max_ = 469.1nm, full width at half maximum (FWHM) = 33nm), then 22 seconds of darkness to assess pupillary redilation before exposure to 9 seconds of exponentially increasing red light (11.9 to 14.3 Log photons/cm^2^/s ; λ_max_ = 640.1nm, FWHM = 17nm), and finally 10 seconds of darkness to assess redilation. The PLR in patients with high myopia and glaucoma was significantly reduced compared to emmetropes and high myopes without glaucoma. Data are presented as mean ± standard error of the mean.

### Data Analysis and Statistics

Pupil size over time were processed for blink artifact removal using a semi-automated software by two independent reviewers (MTF, RPN), and pupil sizes were expressed as percent constriction from baseline pupil size:

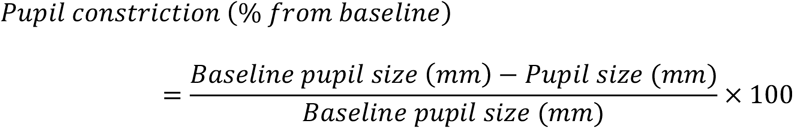

Baseline pupil size was calculated as the median horizontal radius of the pupil during the 5 seconds preceding blue light onset. Normalizing pupil constriction to individual baselines accounted for differences in pupil sizes in darkness between participants, allowing constriction responses to be comparable.

Fifteen pupillometric features, including baseline pupil size, were extracted from the processed pupil constriction traces of each participant (Supplementary Table 1). These features, along with demographic and clinical characteristics of participants, were compared between the EM, HM, and HMG groups using appropriate statistical tests detailed below. For categorical data, differences in sex and cataract status across groups were assessed using a χ^2^ test, and differences in ethnicity were assessed using Fisher’s Exact Test. For continuous variables, normal and homoscedastic data were compared using one-way ANOVA followed by post-hoc Tukey tests. Normal and heteroscedastic data were compared using Welch’s ANOVA and post-hoc Games-Howell tests, while non-normal data were compared using Kruskal-Wallis one-way ANOVA and post-hoc Dunn’s tests. The threshold for significance for all statistical tests was set at α = 0.05, and all post-hoc tests employed Benjamini-Hochberg correction for multiple comparisons.

Logistic regression was used to distinguish glaucomatous high myopes from a combined population of high myopes and emmetropes without glaucoma. Models were generated using combinations of five PLR features found to be associated with glaucoma as linear predictors,^37^ and the model with the greatest area under the receiver operating characteristic curve (AUC) was selected as the final model.^37^ Predictors considered included Phasic Constriction to Blue Light, Maximum Constriction to Blue Light, Post-Illumination Pupillary Response (PIPR) to Blue Light taken as the area under the pupil constriction Curve 0-12 seconds following light offset (PIPR AUC 0-12s), Phasic Constriction to Red Light, and Maximum Constriction to Red Light; interaction terms were not considered to avoid overfitting. The final logistic regression model (LRM) used Phasic Constriction to Blue Light, Maximum Constriction to Blue Light, and Phasic Constriction to Red Light as predictors, and the optimum classification threshold was selected using Youden’s J, with confidence intervals computed using a bootstrap procedure (n=2000) for sensitivity and specificity, and computed using DeLong’s method for the AUC. All statistical analyses were conducted using RStudio version 1.2.1335 running R version 3.6.0: A Language and Environment for Statistical Computing (R Core Team, Vienna, Austria). Figures were generated using the ggplot2 package.^44^

## Results

### Demographic and clinical characteristics of study participants

From the population that took part in this study (n = 73), one participant from the HM group was excluded due to a family history of glaucoma. Three patients were excluded from the HMG group due to dense cataracts (> NS2+; n = 2) or for intraocular lenses (n = 1). The OCT data of one patient from the HMG group could not be collected, but their data was included in other analyses. The final study population comprised 28 emmetropes (age 52.9 [12.5] years, median [IQR], 42.9% male, 85.7% Chinese), 24 high myopes without glaucoma (age 43.9 [19.0] years, median [IQR], 50.0% male, 95.8% Chinese), and 17 high myopes with glaucoma (age 57.4 [13.3] years, median [IQR], 82.4% male, 94.1% Chinese). Within the HMG group, 17 patients were diagnosed with primary open-angle glaucoma (7 with normal tension glaucoma). Groups were not significantly different in age (P=0.09) and ethnicity (P=0.88). Distribution of sex was different across groups (P=0.03), with the HMG group having the greatest proportion of males, but no comparisons retained significance during post-hoc testing. Cataract status was also different between groups (P=0.002), with the HMG group having more cataracts than the EM group. As expected, emmetropes had significantly greater spherical equivalents (SE) than both high myopia groups (P<0.001), who had similar SE. VFMD was also significantly reduced in the HMG group (P<0.001) compared to the EM and HM groups; HM participants had slightly reduced VFMD compared to EM, but post-hoc comparison did not reach significance (P=0.07). Average RNFL thinning was reported in the HMG group compared to both groups (P<0.001). Post-hoc tests also revealed thinner average RNFL thickness in HM compared to EM (P=0.03), in addition to superior (P=0.005) and inferior (P<0.001) quadrant RNFL thinning, and a temporal quadrant RNFL thickening (P=0.006). While we repeated the OCT assessment whenever scans were of unreliable quality, OCT signal strengths were still lower in both HM and HMG groups compared to the EM group (P<0.001). The demographic and clinical characteristics of participants are detailed in Table 1.

**Table 1.**
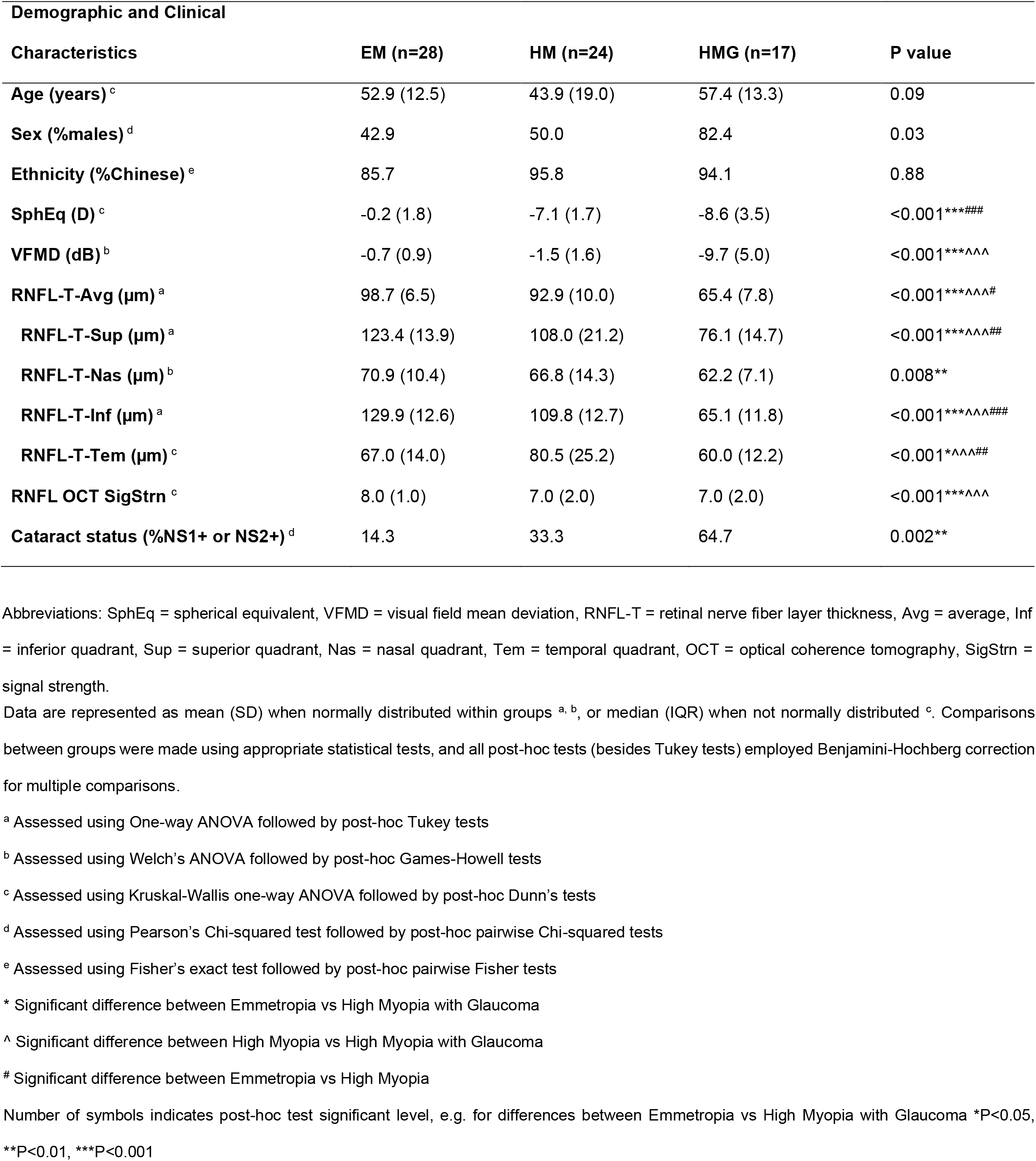
Demographic and clinical characteristics of the study groups.

### Pupillary responses to blue and red light are altered only in eyes with high myopia and glaucoma

Baseline pupil size in darkness was similar across groups (P=0.23) (Figure 2A, Table 2). Shortly after light onset, all eyes exhibited fast phasic pupil constriction, followed by gradual constriction in response to the exponentially increasing blue and red light stimuli (Figure 1). All pupillometric features were similar between EM and HM groups, while multiple pupillometric features were attenuated in the HMG group (Figure 2, Table 2). Principally, phasic constriction to blue (P<0.001) and red (P=0.006) lights, as well as maximum constriction to blue light (P<0.001) were reduced in the HMG compared to the EM and HM groups (Figure 2B, C, F). Maximum constriction to red light was lower in the HMG group than the HM group (P=0.04), and the post-hoc comparison between the HMG and EM groups was marginally significant (P=0.07; Figure 2G). Constriction latency to blue light was also longer (P=0.004) in the HMG group versus the EM and HM groups, while constriction latency to red light was similar across groups (P=0.27).

**Table 2.**
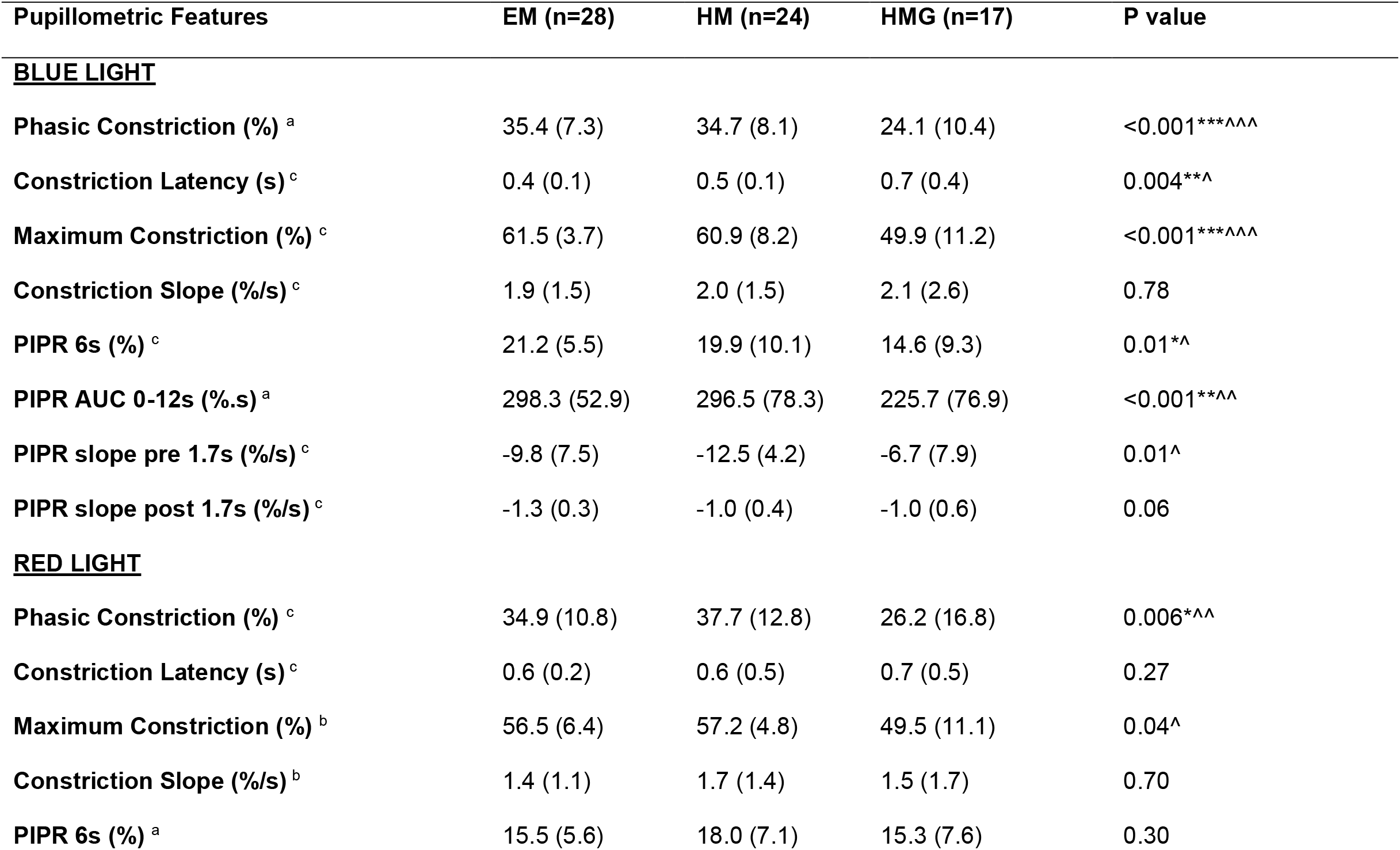
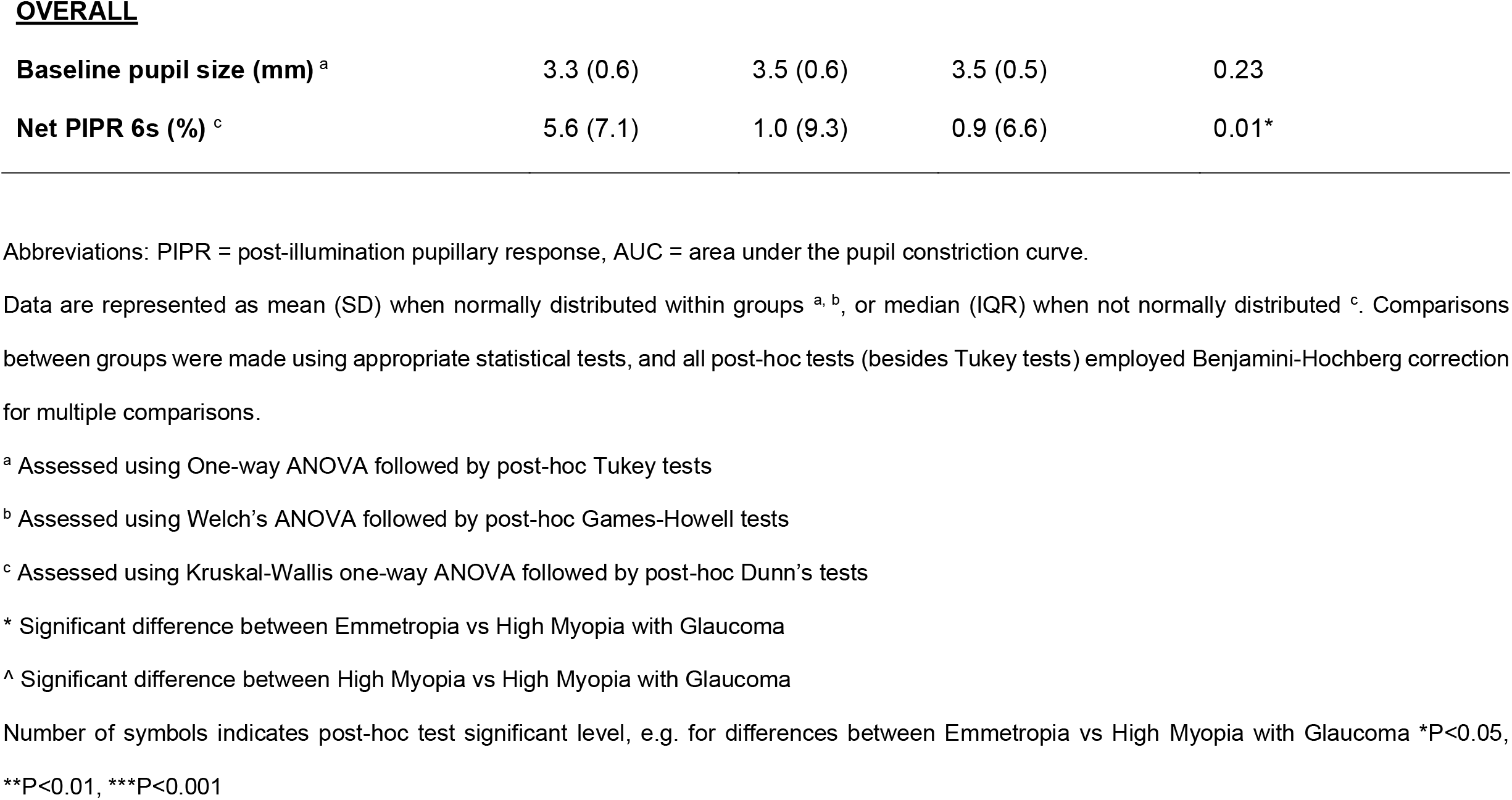
Pupillometric features in emmetropic eyes (EM), eyes with high myopia (HM), and eyes with high myopia with glaucoma (HMG).

**Figure 2.**
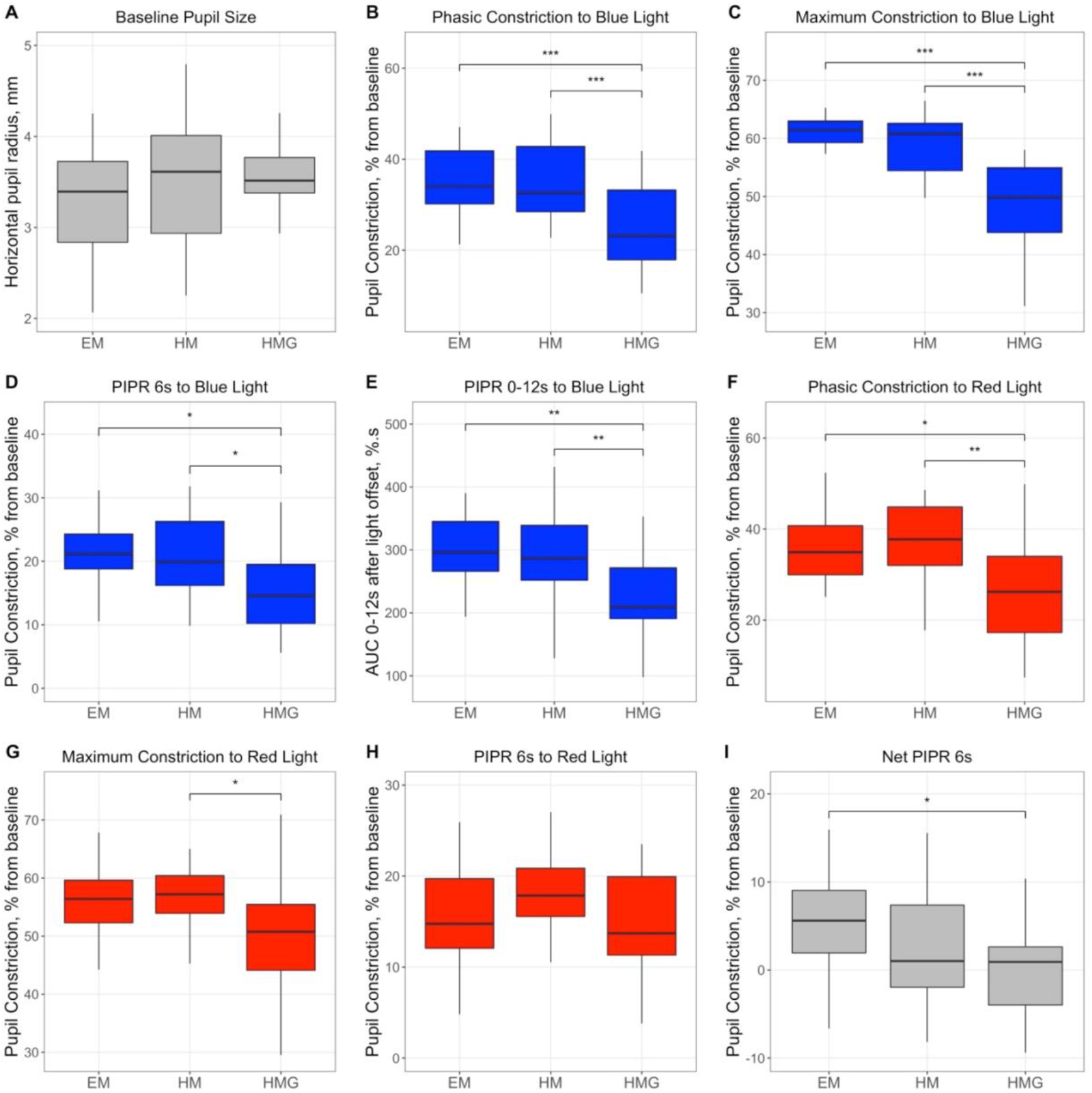
Comparison of predominant pupillometric features between the groups with emmetropia (EM), high myopia (HM) and high myopia and glaucoma (HMG). Data were compared using one-way ANOVA and post-hoc Tukey tests when groups were normally distributed with homogenous variance (**A, B, E, H**), or Welch ANOVA and post-hoc Games-Howell tests with heterogenous variance (**G**), or Kruskal-Wallis one-way ANOVA and post-hoc Dunn’s tests when data were not normally distributed (**C, D, F, I**). Significant differences on post-hoc testing are represented as *P<0.05, **P<0.01, ***P<0.001.

Pupillary redilation following light offset including pupillometric features of melanopsin were also affected in the HMG group. The PIPR to blue light taken as both the area under the redilation curve (PIPR AUC 0-12s) and the median constriction 5 to 7 seconds after light offset (PIPR 6s) were reduced compared to both EM and HM groups (PIPR AUC 0-12s: P<0.001; PIPR 6s: P=0.01) (Figure 2D, E; Table 2). The magnitude of the PIPR slope pre 1.7s was also different between groups (P=0.01) with post-hoc tests showing significant attenuations in the HMG group compared to HM (Table 2). The PIPR 6s to red light was similar across groups (P=0.30), while the Net PIPR 6s was reduced in the HMG group compared to EM (P=0.01) (Figure 2H, I).

### Handheld chromatic pupillometry highlights functional glaucomatous damage in eyes with high myopia

As a proof of concept assessment, pupillometric outcomes were used to distinguish eyes with glaucoma and high myopia (n=17) from eyes without glaucoma (EM and HM), who were shown to have similar PLRs (n=52). LRMs were generated using combinations of five PLR features known to be associated with glaucoma as predictors,^37^ and the model with the highest AUC was selected. The final model used Phasic Constriction to Blue Light, Maximum Constriction to Blue Light, and Phasic Constriction to Red Light as predictors, achieving an AUC of 0.89 (95%CI: 0.77–1.00), with sensitivity 94.1% (95%CI: 82.4%–100.0%) and specificity 78.8% (95%CI: 67.3%–90.4%) (Figure 3). Maximum Constriction to Blue Light was the only significant variable in the final model (P=0.004), with an adjusted odds ratio of 0.81 (95%CI: 0.70–0.94).

**Figure 3.**
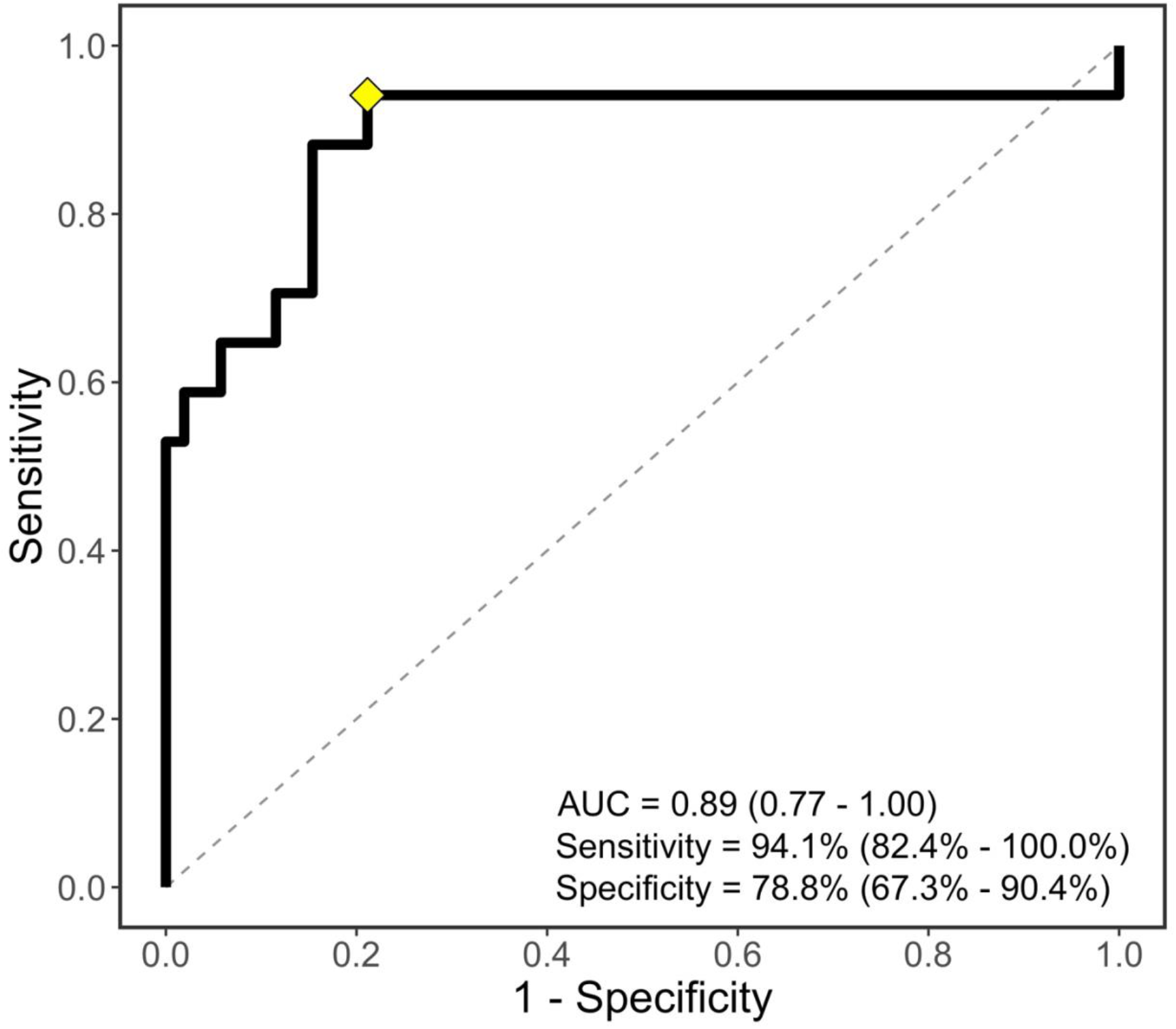
Receiver operating characteristic (ROC) curve for the final logistic regression model for detecting high myopes with glaucoma (n=17) from a combined population of emmetropes and high myopes without glaucoma (n=52). Logistic regression models were generated using combinations of five PLR features known to be associated with glaucoma as predictors, and the model with the greatest Area Under the receiver operating characteristic Curve (AUC) was selected. Interaction terms were not considered to avoid overfitting. The final model used Phasic Constriction to Blue Light, Maximum Constriction to Blue Light, and Phasic Constriction to Red Light as predictors. This model had an AUC of 0.89 (95%CI: 0.77–1.00), and the optimum threshold was selected using Youden’s J with sensitivity 94.1% (95%CI: 82.4%–100.0%) and specificity 78.8% (95%CI: 67.3%– 90.4%), marked on the ROC curve with a yellow diamond.

## Discussion

In this study, we show that pupillary responses to short (9s) and exponentially increasing blue and red light stimuli are not altered in eyes with high myopia in the absence of other diagnosed ocular conditions. Conversely, multiple pupillometric features are affected in glaucomatous eyes with high myopia compared to emmetropic eyes and eyes with high myopia without glaucoma. Using three of these features as predictors in a LRM, HCP can distinguish between eyes with glaucoma from eyes without glaucoma with high accuracy (AUC = 0.89, 95%CI: 0.77–1.00). We have previously demonstrated that both desktop and handheld chromatic pupillometers using exponentially ramping-up light stimulations can detect functional deficits in glaucoma, even at an early stage.^31,33,37^ Using a similar light paradigm, we have also reported that the PLR is unaffected by mild or moderate myopia.^39^ Taken together, our results suggest that HCP using a ramping-up light paradigm can potentially be used to detect glaucoma in populations with a high prevalence of myopia, such as those in East Asia.^2–5^

While decreased functional responses on ERG and SAP have been reported in eyes with high myopia,^9,10,15–19^ prior chromatic pupillometry studies have largely found no myopia-related defects in the PLR.^38–42^ Mutti and colleagues, in contrast, found PLR attenuations in mild to moderate myopia consistent with reduced ipRGC activity.^43^ This is likely due to the author’s unique light paradigm, which stimulated ipRGCs using repeated flashes of blue light, and thus assesses an adaptive change in the PLR rather than basal ipRGC activity assessed following darkness. While none of the aforementioned studies focused on the PLR in eyes with high myopia, our findings agree with the growing body of work that myopia, and even high myopia, does not significantly affect the PLR assessed in darkness. We did, however, observe slight, nonspecific increases in the PLR to red light in the HM versus EM groups, but none of these comparisons reached significance. We have previously reported similar findings in the PLR to red light in eyes with moderate myopia at the high irradiances used in this study, and attributed these differences to confounding longitudinal chromatic aberrations.^39^

On the other hand, eyes with glaucoma and high myopia displayed attenuations in the PLR that agree with retinal ganglion cell dysfunction reported using steady-state pattern ERG in eyes with high myopia and glaucoma,^45–47^ and reverberate established glaucomatous pupillometric alterations.^31–34,37^ The maximum constriction to blue light assessed at high irradiances (14.4 Log photons/cm^2^/s), the PIPR 6s and PIPR AUC 0-12s to blue light, and net PIPR were all reduced in glaucomatous high myopes, indicating ipRGC dysfunction. Furthermore, deficits in phasic constriction responses, constriction latency to blue light, and the rod-dominated early PIPR slope (pre 1.7s) also suggest outer retinal dysfunction;^48^ these findings are in agreement with prior histological and electrophysiology reports showing reduced outer retinal function in glaucoma.^49–51^ Using HCP, we have previously observed reductions in each of the aforementioned PLR variables in glaucomatous eyes in a large study population of eyes with and without refractive errors (n = 322).^37^ In contrast to the previous study, however, we did not find significant deficits in the PIPR slope post 1.7s, the constriction latency to red light, or PIPR 6s to red light in high myopes with glaucoma. We interpret this to be due to a smaller glaucoma effect size in these features; indeed, none of these variables were found to contribute more than 5% to a gradient boosting machine learning model that used PLR features to distinguish glaucomatous eyes from healthy controls.^37^ We also do not exclude the possibility of PLR alterations in the HMG group to be affected by subclinical myopic optic neuropathy, which results from axonal stretching due to the morphological changes of highly myopic eyes.^52^

While high myopia is a risk factor for glaucoma, myopia-related optic nerve and retinal abnormalities can make an accurate glaucoma diagnosis challenging.^20,21,53–56^ Here, we found decreased average RNFL thickness with significant superior and inferior quadrant RNFL thinning, a common sign for glaucoma, in both glaucomatous and non-glaucomatous high myopes.^56,57^ In addition, significant temporal RNFL thickening was seen in non-glaucomatous high myopes compared to emmetropes. These findings are in general agreement with the plethora of studies that have investigated RNFL thickness in high myopes.^10–14^ Kang and colleagues investigated high myopes without chorioretinal atrophy, and speculated these changes in the RNFL thickness profile of high myopes to be caused by uneven growth of the lengthened eye during development.^12^ Multiple studies have also suggested OCT results to be less reliable in high myopes.^10,15–17^ Indeed, we found significantly lower OCT signal strengths in both highly myopic groups compared to emmetropes, even though scanning was repeated up to twice more if results were unreliable. Taken together, while the structural defects present in eyes with high myopia may make glaucoma diagnosis challenging via traditional methods, differences in the PLR of eyes with high myopia in glaucoma may allow HCP to distinguish glaucomatous structural damage from RNFL thinning due to high myopia alone.

Our study has a few limitations. First, as our HM group did not have reduced VFMD compared to the EM group, we cannot generalize our findings to highly myopic eyes with significant visual field defects. Indeed, examination of our study population enabled us to first elucidate the effect on the PLR of high myopia without functional abnormalities, confirmed via four years of follow-up. These results provide the foundations for future investigations that aim to unravel how the PLR is affected by non-glaucomatous high myopia with visual field defects, which presents additional challenges when distinguishing glaucomatous damage. Second, because our proof-of-concept study took place in a clinical setting with an artificially high prevalence of glaucoma, the performance characteristics of HCP may differ if it were to be used to detect glaucoma in other settings, such as in the community. However, for the purpose of aiding the diagnosis of glaucoma in highly myopic patients, the higher pre-test probability of a glaucoma diagnosis may make these performance characteristics more applicable. Finally, due to the relatively small number of participants in each group (EM, HM, and HMG), our findings may require further validation in a larger cohort.

In conclusion, this study demonstrates that highly myopic refractive errors do not affect the PLR to ramping up light stimuli in individuals without other ocular conditions. Conversely, handheld chromatic pupillometry can distinguish glaucomatous damage in highly myopic eyes and may be useful to detect/confirm glaucoma in patients with high myopia, both in the clinic and the community. These findings contribute to the growing evidence supporting handheld chromatic pupillometry as an objective, noninvasive tool for evaluating retinal integrity, particularly when traditional scanning modalities may be confounded by conditions like high myopia. Further clinic- and community-based studies are required to confirm the utility of handheld chromatic pupillometry for the detection of co-existing ocular diseases in myopic populations.

## Data Availability

All data produced in the present study are available upon reasonable request to the authors.

## Other Acknowledgments

We would like to thank the research coordinators in the Glaucoma group at the Singapore Eye Research Institute for recruiting patients for this study.

