## Supplementary Table 1 for "Handheld chromatic pupillometry can reliably detect functional glaucomatous damage in eyes with high myopia"

**Supplementary Table 1.** Definitions of extracted pupillometric features.

| Pupillometric Features | Definition |
| --- | --- |
| Baseline pupil size (mm) | Median horizontal pupil radius assessed in darkness during 5 seconds prior to blue light onset |
| Phasic Constriction (%) | Median of the baseline-adjusted pupil size 0.5 to 2.5  seconds after light onset |
| Constriction Latency (s) | Time required from light onset for the baseline-adjusted pupillary constriction to reach 10% in amplitude |
| Maximum Constriction (%) | Maximum amplitude of constriction at light offset |
| Constriction Slope (%/s) | Slope of gradual pupillary constriction during the last 6 seconds of light exposure |
| PIPR 6s (%) | Median of the baseline-adjusted pupil size 5 to 7 seconds after light offset |
| PIPR AUC 0-12s (%.s) | Area under the pupillary response curve 0 to 12 seconds after blue light offset |
| PIPR slope pre 1.7s (%/s) | Slope of pupil redilation 0 to 1.7 seconds following blue light offset |
| PIPR slope post 1.7s (%/s) | Slope of pupil redilation after 1.7 seconds following blue light offset |
| Net PIPR 6s (%) | PIPR 6s (blue) – PIPR 6s (red) |

Abbreviations: PIPR = post-illumination pupillary response, AUC = area under the pupil constriction curve.
